# Evaluating Eight Retrieval-Augmented Generation (RAG) Large Language Models’ Responses to Clinical Questions: A Comparative Study

**DOI:** 10.64898/2026.08.10.26360108

**Authors:** Poppy A. Krump, Mallory N. Blasingame, Taneya Y. Koonce, Annette M. Williams, Jing Su, Nunzia B. Giuse

## Abstract

**Background:** Large language models (LLMs) that use retrieval-augmented generation (RAG) are increasingly used to answer clinical questions, although the evaluation of these systems remains limited. Building on previous studies conducted by our team, this case report aimed to improve upon this knowledge gap by applying a reusable methodology to compare the performance of eight LLMs that utilize RAG techniques for evidence synthesis.

**Case Presentation:** Eight commercially available RAG LLM tools (OpenEvidence, Undermind, Consensus, SciSpace, Elicit, MediSearch, EvidenceHunt, and Scite) were evaluated using twelve ChatGPT-generated clinical questions on the topics of treatment, etiology, and prognosis. To enable comparison, we prompted ChatGPT to identify all key unique medical concepts from the full set of LLM responses to each question. Concepts were categorized as critical (“must-have”) or non-critical (“nice-to-have”) for answering the clinical question. Experienced information scientists were consulted at each step for their expertise. Descriptive statistics and Kruskal-Wallis tests were used to compare performance across tools and question categories. No significant differences were found among the eight RAG LLMs in their coverage of “must-have” (p=0.95) or “nice-to-have” (p=0.16) key unique medical concepts, and no single tool consistently captured all identified concepts.

**Conclusions:** These findings suggest that RAG LLMs may be supplementary tools for evidence retrieval and synthesis but cannot, at this time, fully replace comprehensive expert review of the medical literature. The evaluation framework presented here may be a useful model for future comparative assessments of rapidly evolving AI evidence synthesis tools.

## Background

While artificial intelligence (AI) has shown preliminary promise as an evidence synthesis tool [1], the rapid pace of developments in AI has made it challenging to properly evaluate, compare, and understand tools available on the market [2]. Despite this lack of knowledge, clinicians are increasingly turning to large language models (LLMs) with clinical questions [3]. According to a May 2026 report from NBC News, OpenEvidence states that almost two-thirds of US physicians (approximately 650,000 doctors) are currently using the platform, which is designed to aid healthcare providers in quickly querying the literature to answer clinical questions [4]. OpenEvidence, like many other LLM tools advertised for clinicians, students, and researchers, uses retrieval-augmented generation (RAG) techniques in response to user queries [5]. Retrieval-augmented generation (RAG) enhances large language model performance by grounding responses in information retrieved from pre-selected external sources [2]. By incorporating RAG techniques, companies developing LLM tools can potentially address concerns about LLM use in evidence synthesis [6], namely by 1) providing stronger guardrails, 2) decreasing hallucinations, and 3) improving timeliness of information [7].

Recent systematic reviews reporting on the implementation of RAG LLMs in healthcare have highlighted uses including medical question-answering, information retrieval from datasets, and clinical decision support [8,9]. These RAG LLM tools can leverage diverse knowledge sources including clinical practice guidelines [10,11,12], electronic health record (EHR) data [13,14], institutional resources such as EHR usage manuals and regulatory guidance documents [15,16], and trusted medical evidence from the biomedical literature [17,18]. Gargari et al.’s narrative review of RAG use across a range of medical domains demonstrates improved accuracy, reliability, and efficiency in LLMs with RAG integration compared to base models alone [19]. However, it is important to note that many of the aforementioned examples of evaluations of RAG LLM tools in healthcare reflect scenarios where custom RAG systems are developed and evaluated in an internal setting for specific users’ needs [15,16]. Furthermore, recent studies have challenged the assumption that RAG improves model performance compared to frontier LLMs – which further complicates the interpretation of evidence regarding RAG LLM performance [20,21].

While there have been evaluations of LLMs in performing specific information-related tasks [22,23,24], comparative evaluations of commercially available RAG LLM tools for evidence synthesis are not well-represented in the literature. This case report aimed to improve upon and address this knowledge gap, not only by comparing eight RAG LLM tools, but also by providing a reusable methodology.

This study represents the next step in a series of studies conducted by the Center for Knowledge Management (CKM) at Vanderbilt Health to evaluate the application of generative artificial intelligence for answering clinical questions. Earlier studies by the team include the evaluation of an internally managed chat tool using GPT-4 in answering questions from an in-house database of clinical evidence requests previously answered by medical librarians [25], and a comparison of the responses from 5 LLM tools (ChatGPT, Gemini, Copilot, DeepSeek, and Grok-3) to the ones from information scientists for 45 AI-generated medical questions [26]. In the current study, the CKM team, recognizing the potential for improvement of the LLMs’ capabilities with RAG techniques in evidence synthesis, opted to examine and compare the performance of eight RAG LLM tools in answering a series of clinical questions.

## Case Presentation

### Methods

The study received a non-human subjects research determination from the Vanderbilt University Medical Center Institutional Review Board (IRB #251074). The reporting of this study follows the Chatbot Assessment Reporting Tool (CHART) guidelines [27,28]; the CHART Methodological Diagram and Checklist can be viewed in Appendix A.

### Selection of RAG LLM Tools

In this case study, eight RAG LLM tools were evaluated: OpenEvidence (https://www.openevidence.com/), Undermind (https://www.undermind.ai/), Consensus (https://consensus.app/), SciSpace (https://scispace.com/), Elicit (https://elicit.com/), MediSearch (https://medisearch.io/), EvidenceHunt (https://evidencehunt.com/), and Scite (https://scite.ai/).

Tools were selected via a thorough review of the literature that examined RAG LLMs in the context of evidence synthesis or librarianship. The articles retrieved from this search [29,30,31,32] provided a basis on which tools were chosen. The following criteria for tool selection were applied: those that 1) utilize retrieval-augmented generation techniques and 2) provide a model primarily trained on scientific content. Wolters Kluwer’s UpToDate Expert AI model was considered as a potential tool to evaluate in this study; however, due to restrictions established by UpToDate on the usage of its data by other AI models, which our methodology required, a decision was made not to include it.

With the exception of OpenEvidence, each RAG LLM tool offered “freemium” access, which provided free limited access to the tool as well as fee-based advanced subscription plans. For the seven freemium tools, we selected the highest-tier subscription plan (often referred to as “premium” or “pro”) available at the time (September 2025), so that the evaluation reflected the most advanced AI model available on each of the platforms.

OpenEvidence is free for clinicians with a National Provider Identifier (NPI). For this evaluation, OpenEvidence provided this study team with courtesy access.

While it was not possible to ascertain the full complement of knowledge sources leveraged by the tools selected for the current study, at the time of this investigation, known knowledgebases included Semantic Scholar, OpenAlex, Google Scholar, PubMed, and, in the cases of OpenEvidence and Scite, databases of full-text journal articles from established partnerships with major medical publishers and societies. Two of the study RAG LLMs (Elicit and Undermind) stated that they extracted data from full text when possible and used the abstract if full text was not available. The other tools did not overtly state whether they consulted the full text or abstract for their answers, nor whether they relied primarily on open access articles. The RAG LLMs included in the study drew upon peer reviewed articles, international health guidelines, and articles from preprint servers in their knowledgebases. We do recognize, though, that much may have changed in the time elapsed since the study was conducted.

### Question Generation

As questions from actual clinical encounters at our institution are proprietary and cannot be used with publicly available LLM tools, for this study, the team prompted ChatGPT to generate twelve clinical questions (using the ChatGPT-4.o mini model as a signed-out user). ChatGPT was instructed to generate four questions within each of the following categories: treatment, etiology, and prognosis. These question categories represent the types of queries most often received via our Center’s clinical evidence provision services [33]. ChatGPT was prompted to create these clinical questions in PICO format (patient, intervention, comparison, outcome). Prompts for question generation followed the COSTAR framework [34] and can be found in Appendix B, along with the full list of twelve questions used for the evaluation. To ensure that the questions provided by ChatGPT were medically plausible, each question was reviewed prior to inclusion, by two experienced information scientists with formal education and training in medicine. Each question was subsequently entered into the eight RAG-based LLM tools by a single information scientist in a new chat session while logged into a personal account.

### Evaluation

All RAG LLM responses to the clinical questions were saved in a REDCap database for analysis [35,36]. The study did not use a “reference standard” for comparison but rather compared the tools against each other. An iteratively developed comparison strategy that combined AI assistance with information scientist review was utilized. For each of the twelve AI-generated clinical questions, the eight responses were consolidated into a single file and fed into ChatGPT. Study authors created a prompt asking ChatGPT to identify all the unique information elements within the consolidated file of eight answers. Once ChatGPT provided the list of unique information elements for each question, we then asked the tool to devise a list of broad medical concepts into which the unique elements could be grouped. The lists of unique medical information elements and key unique medical concepts were both carefully reviewed by the information scientist prompting ChatGPT to ensure that the elements and concepts were correctly representing the RAG LLM tools’ answers. ChatGPT was then asked to identify which of the key unique medical concepts were present in each tool’s individual response to the question. As with the previous step, the responses from ChatGPT were verified to confirm that the key unique medical concepts it identified as included in each tool’s answer were actually represented. Additionally, ChatGPT was asked to determine which of the key unique medical concepts were critical, or “must-have” for answering each question, and which were “nice-to-have,” or not directly related to answering the clinical question. To assess whether a key unique medical concept was truly “must-have,” two experienced information scientists, one of whom also had formal medical training, reviewed the list generated by ChatGPT and made corrections if needed.

All prompts submitted to ChatGPT were developed and tested by a team of information scientists, and the prompts used can be found in Appendix C. All evaluation steps were conducted while logged into a paid ChatGPT business account to enable submission of PDFs to the tool. At the time of the evaluation (March 2026), ChatGPT Business used the GPT-5 model; the ‘Auto’ option (which automatically decides the best processing mode based on the prompt) was used for all conversations.

To illustrate the distinction between unique information elements, key unique medical concepts, and critical (“must-have”) and non-critical (“nice-to-have”) unique medical concepts, consider an etiology question generated for this evaluation: *In pediatric patients presenting with asthma exacerbations, does early-life exposure to household mold, compared with no significant mold exposure, contribute to an increased frequency of exacerbations?* One unique information element identified in the response from Undermind was: “Specific mold species such as Penicillium, Mucor, and Aspergillus are associated with increased asthma morbidity and difficult-to-control asthma, particularly among sensitized children.” This information element was categorized under the broader key unique medical concept “Specific Mold Species and Fungal Exposure in Asthma Morbidity.” Although this concept was considered relevant, both ChatGPT and the information scientist reviewers classified it as a “nice-to-have” concept rather than one that was critical for answering the clinical question. The concept “Dose-Response Relationship Between Mold Exposure and Asthma Morbidity” was identified as a critical, or “must-have,” concept for this question, as it directly addresses the relationship between the extent of mold exposure and the frequency of asthma exacerbations in pediatric patients.

## Data Analysis

Descriptive statistics were used to report on the number of “must-have” key unique medical concepts and “nice-to-have” concepts for each individual question, by question category, and for all twelve questions combined.

Kruskal-Wallis tests were used to assess whether there were any significant differences among the tools in terms of their coverage of the “must-have” key unique medical concepts and of the concepts deemed “nice-to-have.” Kruskal-Wallis testing was also conducted to determine if there were any significant differences between the tools’ coverage of “must-have” key unique medical concepts or of “nice-to-have” key unique medical concepts by question category (i.e., treatment, etiology, and prognosis).

## Results

Ninety-one key unique medical concepts were identified by ChatGPT from the collective answers each RAG LLM generated to the twelve clinical questions and an information scientist confirmed their actual presence. Out of the 91 key unique medical concepts, 42 were labeled by ChatGPT as critical, or “must-have,” with a remainder of 49 labeled as “nice-to-have.” A thorough review by two senior information scientists resulted in the relabeling of three of the 42 “must-have” concepts to “nice-to-have” and eight of the “nice-to-have” concepts to “must-have” concepts, resulting in this final label reclassification of the 91 concepts: 47 “must-have” and 44 “nice-to-have.”

The “must-have” (47) results are as follows: OpenEvidence included 36 (77%), Undermind included 44 (94%), Consensus included 38 (81%), SciSpace included 41 (87%), Elicit included 42 (89%), MediSearch included 38 (81%), EvidenceHunt included 38 (81%), and Scite included 41 (87%). A Kruskal-Wallis test found no significant differences in the number of “must-have” concepts across the eight tools’ answers (p = 0.95).

The “nice-to-have” (44) results are as follows: OpenEvidence included 28 (64%), Undermind included 28 (64%), Consensus included 27 (61%), SciSpace included 38 (86%), Elicit included 32 (73%), MediSearch included 22 (50%), EvidenceHunt included 24 (55%), and Scite included 29 (66%). A Kruskal-Wallis test found no significant differences in the number of “nice-to-have” concepts across the eight tools’ answers (p = 0.16).

For figures documenting the number of “must-have” and “nice-to-have” key unique medical concepts per RAG LLM by question, see Appendix D and Appendix E.

Figures 1 and 2 show the coverage of “must-have” key unique medical concepts and “nice-to-have” concepts for each tool by question category. A Kruskal-Wallis test showed no significant difference in the tools’ coverage of “must-have” key unique medical concepts by question category (p = 0.94), nor did a Kruskal-Wallis test find a significant difference across the tools on their inclusion of “nice-to-have” key unique medical concepts by question category (p = 0.07).

**Figure 1.**
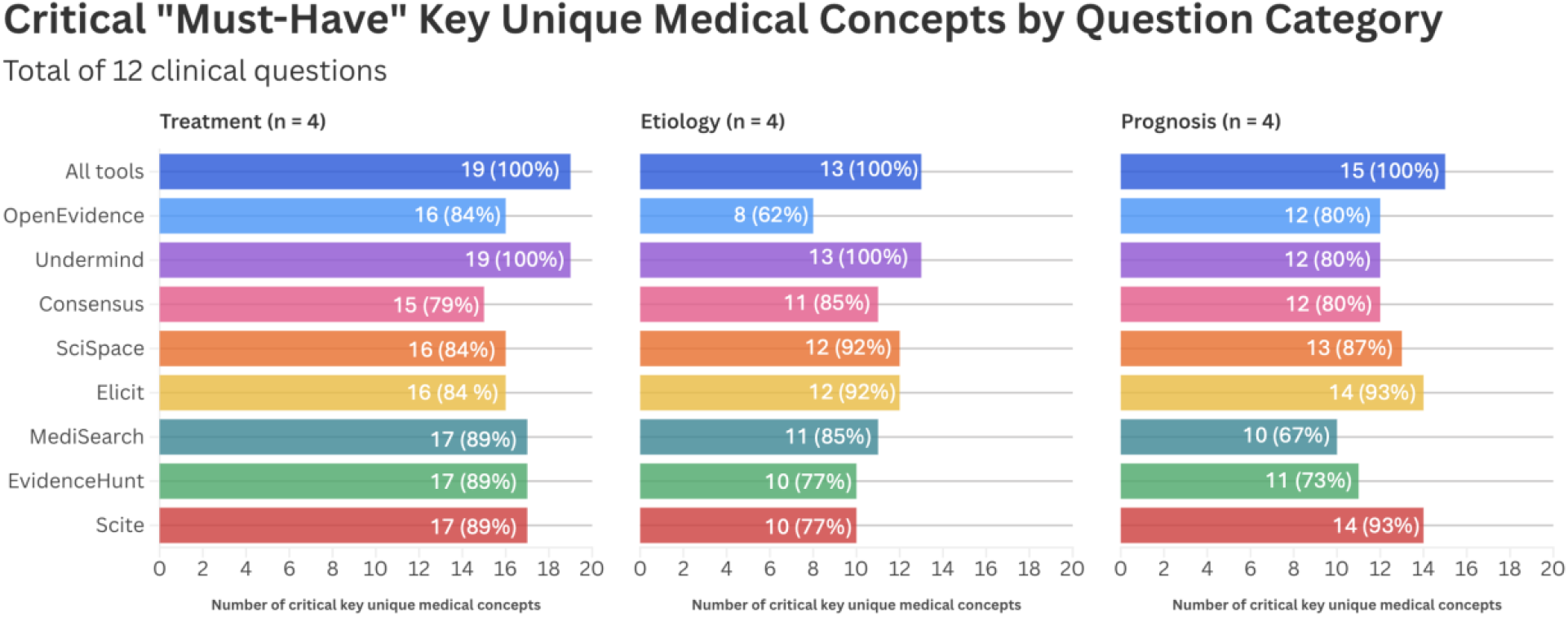

**Figure 2.**
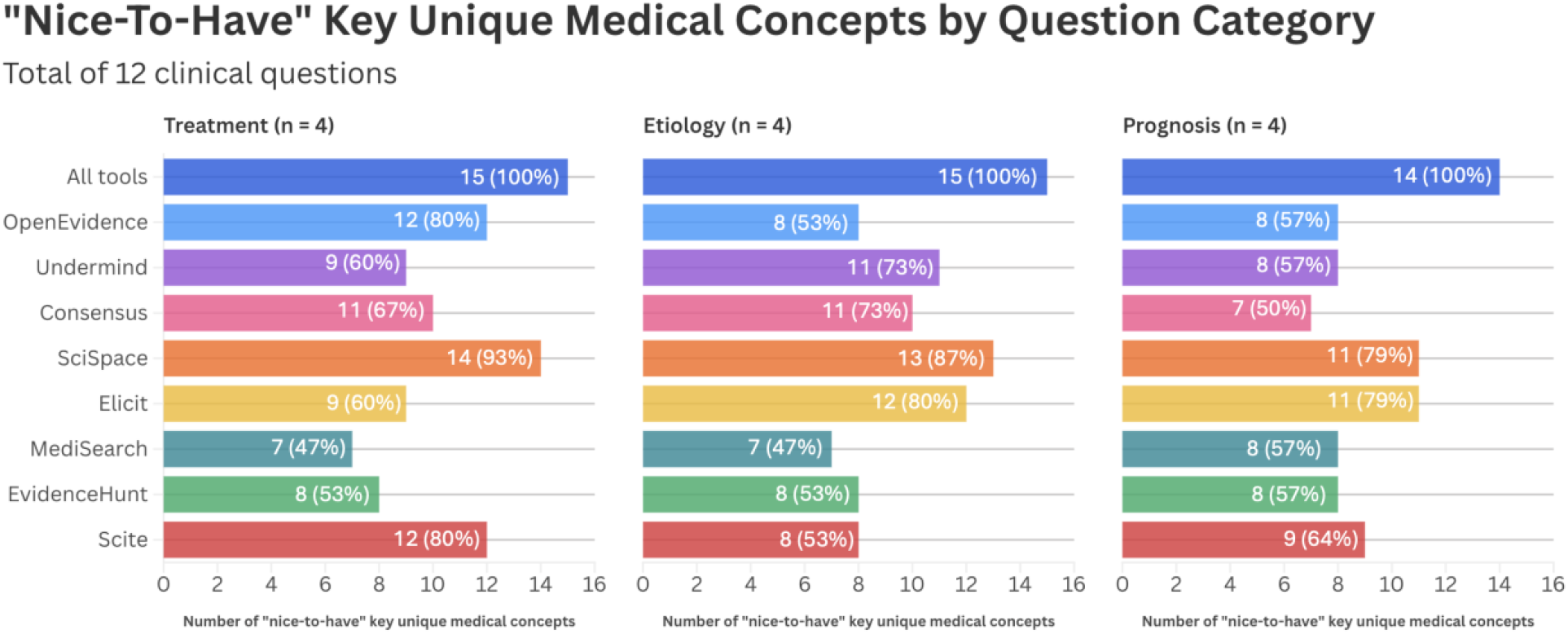

## Discussion

In this comparative evaluation, we found that eight RAG LLMs’ responses to clinical questions contained many of the same medical concepts, but no single tool consistently captured all concepts that were classified as critical, or “must-have,” for answering each question. The lack of complete coverage suggests that evidence syntheses provided by LLMs, even when improved through retrieval-augmented generation, should not, at this time, be considered comprehensive representations of the available evidence. This is consistent with the findings from the Center’s previous studies comparing the responses of LLMs against medical librarians’ answers, which found that both humans and chatbots can include important additional information [26]. The results from this study additionally, align with findings from other evaluations of RAG LLM tools, which emphasize the potential for AI to support, but not replace, human review of medical literature [24].

Although statistical testing did not demonstrate significant differences between tools in their answers, the observed variability suggests that users should nevertheless evaluate each tool based on its capabilities, strengths within specific content domains, and transparency, quality, and appropriateness of sources it consults. Additionally, it is important to emphasize that the absence of statistically significant differences between tools should be interpreted cautiously; given that this was a pilot evaluation, the sample size was not powered for statistical analysis.

This case report did not find any significant difference in the tools’ coverage of “must-have” key unique medical concepts by question category (i.e., treatment, etiology, and prognosis). This is reassuring as studies that investigate clinician information seeking behavior [33,37,38], have found that clinicians tend to ask treatment related questions more often than other question types, which makes it extremely critical to pay attention to how questions in this category are being answered by RAG LLM tools.

The results from this case study add to the literature evaluating the role of retrieval-augmented generation in healthcare. Previous studies have demonstrated that RAG architectures can improve factual accuracy, reduce hallucinations, and increase the relevance of generated responses compared with foundation models alone [2,8,16,17]. However, much of the current published literature has focused on custom-built systems developed for specific institutional or clinical purposes. There is limited research evaluating commercially available RAG LLM tools that are marketed directly to clinicians, students, and researchers. The findings from this study suggest that while these tools may share similar overall capabilities, differences in retrieval strategies and underlying knowledge sources may influence the completeness of responses. Because many commercial tools may not fully disclose their retrieval methods and knowledgebases, users may have difficulty determining why certain information is included or omitted.

In following the ongoing evolution of large language models, we have been able to observe how rapidly the field is advancing. It is a well-known fact that many of the tools are engaging with publishers to expand their content coverage and continuing to refine their models, therefore it is reasonable to imagine that retesting even with the same questions using our methodology would lead to different results. As such, our aim is to aid in promoting awareness and comfort with LLMs in the field of library and information sciences so that we can become expert users and engage in ongoing evaluation, as we are often charged with communicating to our users how these tools can be embedded in their workflows.

This study notably contributes an evaluation framework that combines AI assistance with validation by expert information scientists, allowing for efficient analysis while always maintaining human oversight. This methodology can be easily used by our readership for retesting, as we provide a reusable dataset, including the full list of questions and prompts employed in the study. As the number of commercially available AI evidence tools continues to grow, and as existing tools continue to evolve, robust and replicable evaluation methods such as the approach applied in the current evaluation will be necessary for ongoing LLM performance assessment.

As retrieval-augmented generation becomes increasingly integrated into LLMs used for medical information seeking, thorough evaluation of commercially available tools is essential. Our findings suggest that while current RAG LLMs demonstrate substantial commonality in the concepts they provide, important variation remains, and no single tool consistently captured all concepts. These results support the use of RAG LLMs as supplementary tools for evidence retrieval and synthesis rather than replacements for an expert information scientist’s comprehensive literature search, evaluation, and synthesis of the medical literature.

## AUTHOR CONTRIBUTIONS STATEMENT

Poppy A. Krump: Methodology; investigation; data curation; formal analysis; visualization; writing–original draft; writing–review and editing. Mallory N. Blasingame: Methodology; investigation; data curation; formal analysis; writing–original draft; writing–review and editing. Taneya Y. Koonce: Methodology; investigation; data curation; formal analysis; writing–review and editing. Annette M. Williams: Methodology; investigation; data curation; writing–review and editing. Jing Su: Methodology; investigation; writing–review and editing.

Nunzia B. Giuse: Conceptualization; methodology; investigation; formal analysis; visualization; writing–original draft; writing–review and editing.

## ACKNOWLEDGEMENTS

This research was developed through the training and support provided by the Medical Library Association’s Research Training Institute (RTI).

## DATA AVAILABILITY STATEMENT

All data produced in the present study are available upon reasonable request to the authors.

## References

1. Ruan M, Fan J, Liu M, Meng Z, Zhang X, Zhang C. Artificial intelligence for the science of evidence synthesis: how good are AI-powered tools for automatic literature screening? BMC Med Res Methodol. 2025 Aug 25;25(1):199. DOI: 10.1186/s12874-025-02644-9.

2. Harasgama S, Pearce H, Loftus L, Painter H, Ford J. An overview of artificial intelligence approaches for automating evidence synthesis. Public Health. 2026 May;254:106220. DOI: 10.1016/j.puhe.2026.106220.

3. Center for Digital Health and AI. 2026 Physician Survey on Augmented Intelligence [Internet]. American Medical Association; [cited 10 Aug 2026]. <https://www.ama-assn.org/practice-management/digital-health/physician-survey-augmented-intelligence>.

4. Perlo J. Most U.S. doctors are quietly using this AI tool. Few patients know about it. NBC News [Internet]. 2026 May 13 [cited 10 Aug 2026]. <https://www.nbcnews.com/tech/tech-news/openevidence-ai-doctor-medical-physician-login-app-what-npi-uptodate-rcna341064>.

5. Low YS, Jackson ML, Hyde RJ, et al. Answering real-world clinical questions using large language model, retrieval-augmented generation, and agentic systems. Digit Health. 2025;11:20552076251348850. DOI: 10.1177/20552076251348850.

6. Gartlehner G, Kahwati L, Nussbaumer-Streit B, Crotty K, Hilscher R, Kugley S, et al. From promise to practice: challenges and pitfalls in the evaluation of large language models for data extraction in evidence synthesis. BMJ Evid-Based Med. 2025 Dec 1;30(6):385– 9. DOI: 10.1136/bmjebm-2024-113199.

7. Church KW, Sun J, Yue R, Vickers P, Saba W, Chandrasekar R. Emerging trends: a gentle introduction to RAG. Nat Lang Eng. 2024 Jul;30(4):870–81. DOI: 10.1017/S1351324924000044.

8. Liu S, McCoy AB, Wright A. Improving large language model applications in biomedicine with retrieval-augmented generation: a systematic review, meta-analysis, and clinical development guidelines. J Am Med Inform Assoc. 2025 Apr 1;32(4):605–15. DOI: 10.1093/jamia/ocaf008.

9. Amugongo LM, Mascheroni P, Brooks S, Doering S, Seidel J. Retrieval augmented generation for large language models in healthcare: A systematic review. PLOS Digit Health. 2025 Jun;4(6):e0000877. DOI: 10.1371/journal.pdig.0000877.

10. Noll R, Windschmitt J, Hofmann E, Bergmann N, Schaaf J. Retrieval-augmented generation for medical decision-making in emergency care. Annu Int Conf IEEE Eng Med Biol Soc IEEE Eng Med Biol Soc Annu Int Conf. 2025 Jul;2025:1–7. DOI: 10.1109/EMBC58623.2025.11253463.

11. Ke YH, Jin L, Elangovan K, Abdullah HR, Liu N, Sia ATH, et al. Retrieval augmented generation for 10 large language models and its generalizability in assessing medical fitness. NPJ Digit Med. 2025 Apr 5;8(1):187. DOI: 10.1038/s41746-025-01519-z.

12. Salahi-Niri A, Safavi-Naini SAA, Devi J, Naderi N, Sebastian S, Adamina M, et al. Using large language models to integrate international IBD guidelines: A retrieval-augmented generation approach. Colorectal Dis Off J Assoc Coloproctology G B Irel. 2026 Apr;28(4):e70436. DOI: 10.1111/codi.70436.

13. Alkhalaf M, Yu P, Yin M, Deng C. Applying generative AI with retrieval augmented generation to summarize and extract key clinical information from electronic health records. J Biomed Inform. 2024 Aug;156:104662. DOI: 10.1016/j.jbi.2024.104662.

14. Lopez I, Swaminathan A, Vedula K, Narayanan S, Nateghi Haredasht F, Ma SP, et al. Clinical entity augmented retrieval for clinical information extraction. NPJ Digit Med. 2025 Jan 19;8(1):45. DOI: 10.1038/s41746-024-01377-1.

15. Son N, Kang I, Kim I, Lee K, Nam S, Lee D. Development and evaluation of a retrieval-augmented generation-based electronic medical record chatbot system. Healthc Inform Res. 2025 Jul;31(3):218–25. DOI: 10.4258/hir.2025.31.3.218.

16. Nanua S, Steward R, Neely B, Datto M, Youens K. Retrieval-augmented generation for interpreting clinical laboratory regulations using large language models. J Pathol Inform. 2025 Nov;19:100520. DOI: 10.1016/j.jpi.2025.100520.

17. Pearson S, Reyburn M, Foley C, Finch A, Bench S, Bonnici T, et al. Retrieval-augmented generation versus GPT-4o for patient-facing gynecological cancer information: quality evaluation. JMIR Form Res. 2026 Apr 24;10:e90139. DOI: 10.2196/90139.

18. Li Y, Du X, Wang Y, Chen X, Zhou Z, Lian J, et al. AI-assisted literature screening: a hybrid approach using large language models and retrieval-augmented generation. Int J Med Inf. 2026 Mar 1;207:106205. DOI: 10.1016/j.ijmedinf.2025.106205.

19. Gargari OK, Habibi G. Enhancing medical AI with retrieval-augmented generation: a mini narrative review. Digit Health. 2025;11:20552076251337177. DOI: 10.1177/20552076251337177.

20. Vishwanath K, Alyakin A, Ghosh M, et al. General-purpose large language models outperform specialized clinical AI tools on medical benchmarks. Nat Med. 2026;32(7):2405–2409. DOI: 10.1038/s41591-026-04431-5.

21. Jung H, Cho K, Jun TJ, Kim YH. RAG in clinical practice: a cautionary tale of AI ‘Truthfulness.’ Npj Health Syst. 2026;3:57. DOI: 10.1038/s44401-026-00115-x.

22. Labenbacher S, Niederer M, Hammer S, Bader M, Schreiber N, Bornemann-Cimenti H. Performance of AI tools in citing retracted literature: content analysis. J Med Internet Res. 2026 May 1;28:e88766. DOI: 10.2196/88766.

23. Raposio E, Baldelli I. Reliability and accuracy of generative artificial intelligence tools in providing general information on migraine surgery. Plast Reconstr Surg Glob Open. 2025 Oct;13(10):e7176. DOI: 10.1097/GOX.0000000000007176.

24. O’Rourke J, Byrne M, Schroers G. Artificial intelligence applications versus manual methods for literature retrieval: a comparative analysis. West J Nurs Res. 2026 Jun 10;1939459261451723. DOI: 10.1177/01939459261451723.

25. Blasingame MN, Koonce TY, Williams AM, Giuse DA, Su J, Krump PA, Giuse NB. Evaluating a large language model’s ability to answer clinicians’ requests for evidence summaries. J Med Libr Assoc JMLA. 2025 Jan 14;113(1):65–77. DOI: 10.5195/jmla.2025.1985.

26. Blasingame MN, Koonce TY, Williams AM, Su J, Giuse DA, Krump PA, Giuse NB. Comparing five generative AI chatbots’ answers to LLM-generated clinical questions with medical information scientists’ evidence summaries. J Med Libr Assoc JMLA. 2026 Apr 1;114(2):94–104. DOI: 10.5195/jmla.2026.2333.

27. CHART Collaborative. Reporting guidelines for chatbot health advice studies: explanation and elaboration for the Chatbot Assessment Reporting Tool (CHART). BMJ. 2025 Aug 1;390:e083305. DOI: 10.1136/bmj-2024-083305.

28. CHART Collaborative, Huo B, Collins GS, Chartash D, Thirunavukarasu AJ, Flanagin A, et al. Reporting Guideline for Chatbot Health Advice Studies: The CHART Statement. JAMA Netw Open. 2025 Aug 1;8(8):e2530220. DOI: 10.1001/jamanetworkopen.2025.30220.

29. Westrick J, Juarez L, Hilton S. PubMed vs artificial intelligence: comparison of search results [Internet]. Medical Library Association; 2025 [cited 10 Aug 2026]. <https://library.rush.edu/librarian-scholarly-research/mla-2025-ai>.

30. AkpInar H. Comparison of responses from different artificial intelligence-powered chatbots regarding the All-on-four dental implant concept. BMC Oral Health. 2025 Jun 5;25(1):922. DOI: 10.1186/s12903-025-06294-7.

31. Giglio AD, da Costa MUP. The use of artificial intelligence to improve the scientific writing of non-native English speakers. Rev Assoc Med Bras. 2023;69(9):e20230560. DOI: 10.1590/1806-9282.20230560.

32. Van IJzendoorn DGP, Habets PC, Vinkers CH, Otte WM. Clinical study type classification, validation, and PubMed filter comparison with natural language processing and active learning. medRxiv [Preprint]; 2022 [cited 10 Aug 2026]. Available from: http://medrxiv.org/lookup/doi/10.1101/2022.11.01.22281685.

33. Jerome RN, Giuse NB, Gish KW, Sathe NA, Dietrich MS. Information needs of clinical teams: analysis of questions received by the Clinical Informatics Consult Service. Bull Med Libr Assoc. 2001 Apr;89(2):177–84.

34. GovTech Data Science & AI Division. Prompt engineering playbook (Beta v3) [Internet]. Government of Singapore; 30 Aug 2023 [cited 10 Aug 2026]. <https://www.developer.tech.gov.sg/products/collections/data-science-and-artificial-intelligence/playbooks/prompt-engineering-playbook-beta-v3.pdf>.

35. Harris PA, Taylor R, Thielke R, Payne J, Gonzalez N, Conde JG. Research electronic data capture (REDCap)--a metadata-driven methodology and workflow process for providing translational research informatics support. J Biomed Inform. 2009 Apr;42(2):377–81. DOI: 10.1016/j.jbi.2008.08.010.

36. Harris PA, Taylor R, Minor BL, Elliott V, Fernandez M, O’Neal L, et al. The REDCap consortium: building an international community of software platform partners. J Biomed Inform. 2019 Jul;95:103208. DOI: 10.1016/j.jbi.2019.103208.

37. Ostropolets A, Chen R, Zhang L, Hripcsak G. Characterizing physicians’ information needs related to a gap in knowledge unmet by current evidence. JAMIA Open. 2020 Jul;3(2):281–9. DOI: 10.1093/jamiaopen/ooaa012.

38. Cheng GYT. A study of clinical questions posed by hospital clinicians. J Med Libr Assoc JMLA. 2004 Oct;92(4):445–58.

