## Appendix A for "Evaluating Eight Retrieval-Augmented Generation (RAG) Large Language Models’ Responses to Clinical Questions: A Comparative Study"

**Appendix A. CHART checklist**

| HEADING | # | CHART CHECKLIST ITEM | Page #* |
|---|---|---|---|
| **Title & Abstract** | | | |
| **Title** | **1a** | State that the study is assessing one or more generative AI-driven chatbots for clinical evidence or health advice. | 1 |
| **Abstract/Summary** | **1b** | Apply a structured format, if applicable. | 1 |
| **Introduction** | | | |
| **Background** | **2a** | State the scientific background, rationale, and healthcare context for evaluating the generative AI-driven chatbot(s), referencing relevant literature when applicable. | 1-2 |
| | **2b** | State the aims and research questions including the target audience, intervention, comparator(s), and outcome(s). | 2-3 |
| **Methods** | | | |
| **Model Identifiers** | **3a** | State the name and version identifier(s) of the generative AI model(s) and chatbot(s) under evaluation, as well as their date of release or last update. | 3-4 |
| | **3b** | State whether the generative AI model(s) and chatbot(s) are open-source or closed-source/proprietary. | 3 |
| **Model Details** | **4a** | State whether the generative AI model was a base model or a novel base model, tuned model, or fine-tuned model. | N/A – all tools evaluated were closed-source/proprietary |
| | **4b** | If a base model is used, cite its development in sufficient detail to identify the model. | N/A |
| | **4c** | If a novel base model, tuned model, or fine-tuned model is used, describe the pre- and/or post-implementation/deployment data and parameters. | N/A |
| **Prompt Engineering** | **5a** | Describe the evolution of study prompt development. | 4-5, Appendixes B and C |
| | **5ai** | Describe the sources of prompts. | 3-4 |
| | **5aii** | State the number and characteristics of the individual(s) involved in prompt engineering. | 4 |
| | **5aiii** | Provide details of any patient and public involvement during prompt engineering. | N/A |
| | **5b** | Provide study prompts. | Appendixes B and C |
| **Query Strategy** | **6a** | State route of access to generative AI model. | 3-4 |
| | **6b** | State the date(s) and location(s) of queries for the generative AI-driven chatbot(s) including the day, month, and year as well as city and country. | Appendix A |
| | **6c** | Describe whether prompts were input into separate chat session(s). | 3-4, Appendix C |

| | | | |
|---|---|---|---|
| | **6d** | Provide all generative AI-driven chatbot output/responses | 10 (via Data Availability Statement) |
| **Performance Evaluation** | **7a** | Define the ground truth or reference standard used to define successful generative AI-driven chatbot performance. | N/A – the study did not use a "reference standard." This is described on page 4. |
| | **7b** | Describe the process undertaken for generative AI-driven chatbot performance evaluation. | 4-5 |
| | **7bi** | State the number and characteristics of team members involved in performance evaluation. | 4-5 |
| | **7bii** | Provide details of any patients and public involvement during the evaluation process. | N/A |
| | **7biii** | State whether evaluators were blinded to the identity of the generative AI-driven chatbot(s) under assessment. | 5 |
| **Sample Size** | **8** | Report how the sample size was determined. | 8 |
| **Data Analysis** | **9a** | Describe statistical analysis methods, including any evaluation of reproducibility of generative AI-driven chatbot responses. | 6 |
| | **9ai** | Report the measures used for performance evaluation. | 4-6 |
| **Results** | | | |
| | **10a** | Report the alignment between generative AI-driven chatbot output and ground truth or reference standard using quantitative or mixed methods approaches as applicable. | N/A – no ground truth or reference standard used in this study |
| | **10b** | For responses deviating from the ground truth or reference standard, state the nature of the difference(s). | N/A – no ground truth or reference standard used in this study |
| | **10c** | Report the assessment for potentially harmful, biased, or misleading responses. | N/A – assessment for potentially harmful, biased, or misleading responses not conducted in this case study |
| **Discussion** | | | |
| | **11a** | Interpret study findings in the context of relevant evidence. | 8-9 |
| | **11b** | Describe the strengths and limitations of the study. | 8-9 |

| | | | |
|---|---|---|---|
| | **11c** | Describe the potential implications for practice, education, policy, regulation, and research. | 8-9 |
| **Open Science** | | | |
| **Disclosures** | **12a** | Report any relevant conflicts of interest for all authors. | 10 |
| **Funding** | **12b** | Report sources of funding and their role in the conduct and reporting of the study. | 10 |
| **Ethics** | **12c** | Describe the process undertaken for ethical approval. | 3 |
| | **12ci** | Describe the measures taken to safeguard data privacy of patient health information, as applicable. | N/A |
| | **12cii** | State whether permission/licensing was obtained for the use of original, copyrighted data. | N/A |
| **Protocol** | **12d** | Provide a study protocol. | N/A |
| **Data availability** | **12e** | State where study data, code repository, and model parameters can be accessed. | 10 |

*If in supplementary appendix, indicate "supp" and appendix #, if applicable.

*From:* The CHART Collaborative; Huo B, Collins GS, Chartash D, Thirunavukarasu AJ, Flanagin A, et al. Reporting guideline for chatbot health advice studies: the CHART statement. JAMA Netw Open. 2025 Aug 1;8(8):e2530220. DOI: https://doi.org/10.1001/jamanetworkopen.2025.30220

CHART Methodological Diagram

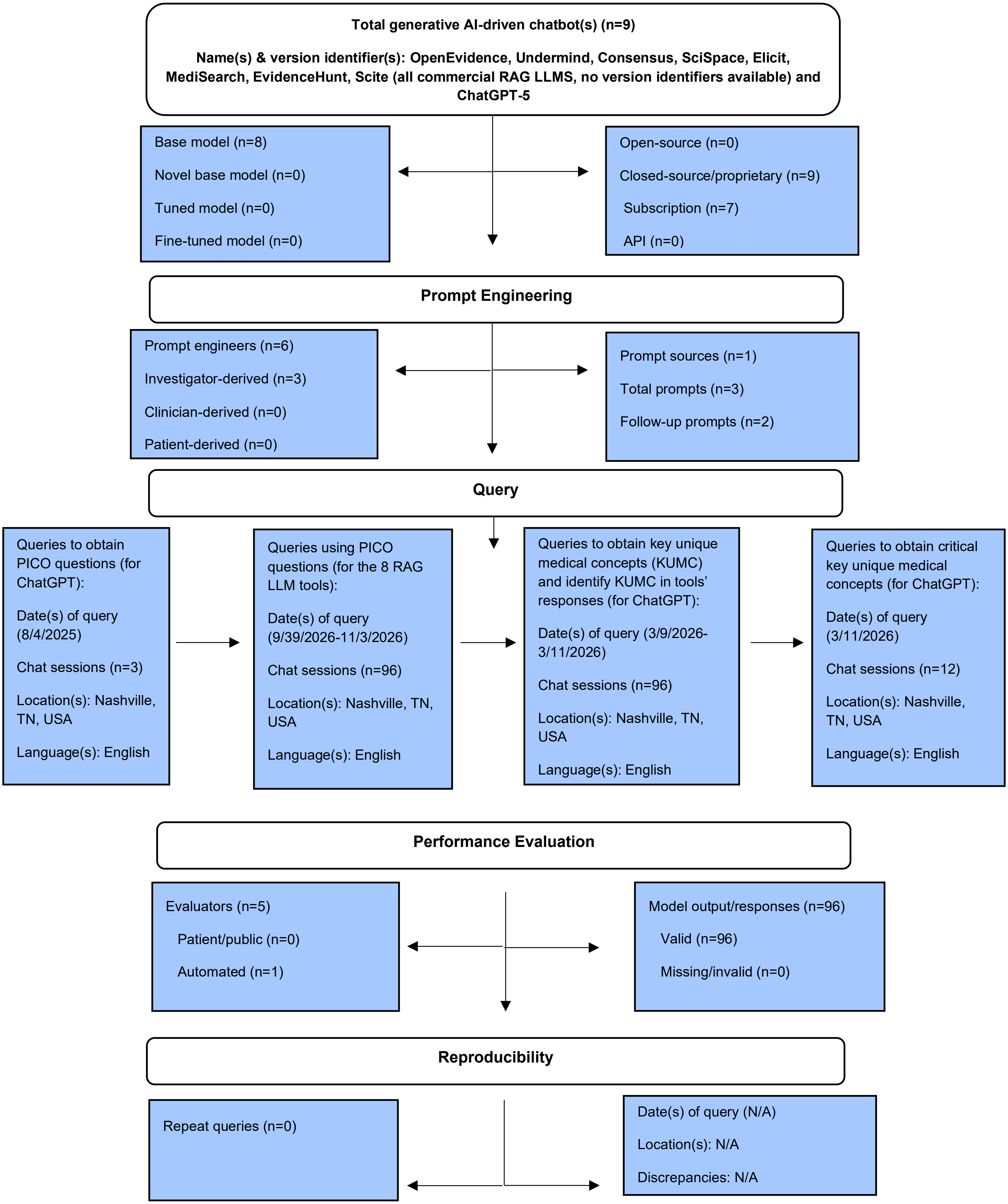
