## Appendix B for "Evaluating Eight Retrieval-Augmented Generation (RAG) Large Language Models’ Responses to Clinical Questions: A Comparative Study"

**Appendix B. List of PICO questions and prompts used for question generation**

*All questions were generated using ChatGPT.*

| Question Number | Question | Question Category |
|---|---|---|
| 1 | In adults hospitalized with community-acquired pneumonia, does adjunctive corticosteroid therapy improve clinical recovery compared to standard antibiotic treatment alone? | Treatment |
| 2 | In patients with newly diagnosed type 2 diabetes, does initiating combination therapy with metformin and a GLP-1 receptor agonist result in better glycemic control compared to starting metformin monotherapy? | Treatment |
| 3 | In elderly patients with atrial fibrillation and a high bleeding risk, does apixaban reduce the incidence of stroke and major bleeding events compared to warfarin? | Treatment |
| 4 | In children with moderate to severe atopic dermatitis, does treatment with dupilumab lead to better symptom control and quality of life than topical corticosteroids alone? | Treatment |
| 5 | In adults with new-onset atrial fibrillation, does untreated hyperthyroidism compared to normal thyroid function increase the risk of arrhythmia recurrence after cardioversion? | Etiology |
| 6 | In pediatric patients presenting with asthma exacerbations, does early-life exposure to household mold versus no significant mold exposure contribute to increased frequency of exacerbations? | Etiology |
| 7 | Among immunocompetent adults with community-acquired pneumonia, does recent use of proton pump inhibitors compared to no PPI use increase the likelihood of infection with drug-resistant organisms? | Etiology |
| 8 | In elderly patients with cognitive decline, does a history of mid-life hypertension compared to normotension increase the risk of developing vascular dementia? | Etiology |
| 9 | In adult patients with newly diagnosed stage II colon cancer, how does the presence of microsatellite instability-high (MSI-H) status compared to microsatellite stable (MSS) status affect the risk of disease recurrence within five years? | Prognosis |
| 10 | Among patients hospitalized with acute decompensated heart failure with preserved ejection fraction (HFpEF), how does elevated NT-proBNP on admission compare to normal levels in predicting 30-day readmission rates? | Prognosis |
| 11 | In elderly patients with newly diagnosed Parkinson's disease, how does the presence of REM sleep behavior disorder compared to its absence influence progression to cognitive impairment over a three-year period? | Prognosis |

| | | |
|---|---|---|
| 12 | For patients with biopsy-confirmed non-alcoholic steatohepatitis (NASH), how does advanced fibrosis on liver elastography compared to no or mild fibrosis affect the likelihood of progression to cirrhosis within five years? | Prognosis |

**Prompt 1a**. *Treatment questions*

#CONTEXT#

I am a medical librarian at a major academic health sciences center. In my team, our members provide evidence-based filtered summaries of the biomedical literature for use in patient care.

#OBJECTIVE#

Your task is to generate a list of four clinical questions that a physician might have while taking care of patients. The questions should be in PICO (Patient, Intervention, Comparison, Outcome) format. These questions should address the category of Treatment.

#STYLE#

The questions should be written in a style likely to be used by a practicing physician.

#TONE#

Maintain a professional and direct tone.

#AUDIENCE#

The audience of the questions is a medical librarian who will answer each inquiry with a search of the literature and filtered evidence summary. Assume the clinical librarian is experienced and familiar with the medical topic at hand.

#RESPONSE FORMAT#

Do not individually separate the PICO elements but rather provide a question that seamlessly includes its elements. Additionally, clearly differentiate each of the four questions you provide.

**Prompt 1b**. *Etiology questions*

#CONTEXT#

I am a medical librarian at a major academic health sciences center. In my team, our members provide evidence-based filtered summaries of the biomedical literature for use in patient care.

#OBJECTIVE#

Your task is to generate a list of four clinical questions that a physician might have while taking care of patients. The questions should be in PICO (Patient, Intervention, Comparison, Outcome) format. These questions should address the category of Disease Etiology, defined as “the cause or causes of a disease.”

#STYLE#

The questions should be written in a style likely to be used by a practicing physician.

#TONE#

Maintain a professional and direct tone.

#AUDIENCE#

The audience of the questions is a medical librarian who will answer each inquiry with a search of the literature and filtered evidence summary. Assume the clinical librarian is experienced and familiar with the medical topic at hand.

#RESPONSE FORMAT#

Do not individually separate the PICO elements but rather provide a question that seamlessly includes its elements. Additionally, clearly differentiate each of the five questions you provide.

*Note*: The definition for etiology was taken from MedlinePlus [1].

**Prompt 1c**. *Prognosis questions*

#CONTEXT#

I am a medical librarian at a major academic health sciences center. In my team, our members provide evidence-based filtered summaries of the biomedical literature for use in patient care.

#OBJECTIVE#

Your task is to generate a list of four clinical questions that a physician might have while taking care of patients. The questions should be in PICO (Patient, Intervention, Comparison, Outcome) format. These questions should address the category of Disease Prognosis, defined as "likely outcome or course of a disease; the chance of recovery or recurrence." In the context of Disease Prognosis, the intervention does not have to be a treatment but could be a lab result, EKG/imaging, or other indicator.

#STYLE#

The questions should be written in a style likely to be used by a practicing physician.

#TONE#

Maintain a professional and direct tone.

#AUDIENCE#

The audience of the questions is a medical librarian who will answer each inquiry with a search of the literature and filtered evidence summary. Assume the clinical librarian is experienced and familiar with the medical topic at hand.

#RESPONSE FORMAT#

Do not individually separate the PICO elements but rather provide a question that seamlessly includes its elements. Additionally, clearly differentiate each of the five questions you provide.

*Note*: The definition for prognosis was taken from the National Cancer Institute [2].

*All prompts were adapted from a prompt used in our team's previous study [3], which was developed based on the COSTAR framework [4].*

**References:**


1. A.D.A.M. Medical Encyclopedia [Internet]. Johns Creek (GA): Ebix, Inc., A.D.A.M.; c1997-2025. Etiology; [updated 8 Jul 2025; reviewed 1 Apr; cited 24 Sept 2025]. <https://medlineplus.gov/ency/article/002356.htm>.
2. National Cancer Institute (NCI) Dictionary of Cancer Terms [Internet]. Bethesda (MD): NCI. Prognosis [accessed 9 Sept 2025]. <https://www.cancer.gov/publications/dictionaries/cancer-terms/def/prognosis>.
3. Blasingame MN, Koonce TY, Williams AM, Giuse DA, Su J, Krump PA, Giuse NB. Evaluating a large language model's ability to answer clinicians' requests for evidence summaries. J Med Libr Assoc. 2025 Jan 14;113(1):65-77. DOI: https://doi.org/10.5195/jmla.2025.1985.

4. GovTech Data Science & AI Division. Prompt engineering playbook (Beta v3) [Internet]. Singapore Government Developer Portal. 2023 [cited 24 Sept 2025]. <https://www.developer.tech.gov.sg/products/collections/data-science-and-artificial-intelligence/playbooks/prompt-engineering-playbook-beta-v3.pdf>.
