## Appendix C for "Evaluating Eight Retrieval-Augmented Generation (RAG) Large Language Models’ Responses to Clinical Questions: A Comparative Study"

**Appendix C. Prompts for evaluation**

**Prompt 2a.** For identifying the unique medical information elements

#CONTEXT# Understand retrieval-augmented generation (RAG) large language model tools' comprehensiveness in answering clinical questions by comparing each tool's performance with a 'summative answer' derived from the fusion of all key clinical concepts expressed at least once in any of the answers generated by the tools used in the study.

#OBJECTIVE# I will provide a clinical question and a PDF document that includes answers generated by eight different large-language models. Without changing any wording included in the PDF document give me the list of the unique medical information elements included.

#STYLE# Write any additional comments you wish to give me in a professional, educated style which emulates the language traditionally used by information scientists/medical librarians while interacting with clinical teams.

#TONE# Maintain a balanced and objective tone.

#AUDIENCE# The target audience is clinicians providing health care.

#RESPONSE FORMAT# A numbered list can be used when more than one unique information element is identified.

#START ANALYSIS# If you understand, ask me to enter the clinical question and PDF document with the answers.

*[input the clinical question and PDF of all 8 answers when prompted]*

**Prompt 2b**. To create the "key unique medical concepts"

Now group what you just gave me into key unique medical concepts.

**Prompt 2c.** To check how many of the "key unique medical concepts" are in each tool's response

*[first branch into a new conversation for each tool]*

Now tell me in a list which of the unique medical information elements, and key unique medical concepts you listed above are included in the uploaded PDF containing the answer of my first* RAG LLM tool. Keep the same identical wording you gave me in the response to the first two prompts. And show me both 1) the exact part of the text that includes the key unique medical concepts, and 2) the exact part of the text that includes each information elements outlined under the key unique medical concepts.

*[ upload the PDF of the tool's response]*

> * this word was updated each time to reflect the number of the tool (second, third, etc.)

**Prompt 3.** To determine which key unique medical concepts are critical

#CONTEXT# To understand retrieval-augmented generation (RAG) large language model tools' comprehensiveness in answering clinical questions we used an LLM to compare each tool's performance with a 'summative answer' represented by a numbered list of key clinical concepts each including unique medical information elements expressed at least once and included under a key unique medical concept. We now want to understand which of the key unique medical concepts ought to be considered critical to answer the clinical question.

#OBJECTIVE# I will provide a clinical question and its numbered list of the key unique medical concepts judged necessary to answer the clinical question. Use the exact wording you are being provided, do not make any changes.

#STYLE# Professional, academic

#TONE# Maintain a balanced and objective tone.

#AUDIENCE# The target audience is clinicians in an academic medical center.

#RESPENCE FORMAT# Number the list provided.

#START ANALYSIS# If you understand, ask me to enter the clinical question and key unique medical concepts which constitute the answer to the question.

*[input the clinical question and related key unique medical concepts]*

<u>*Prompts were developed based on the COSTAR framework [1].*</u>

**References:**


1. GovTech Data Science & AI Division. Prompt engineering playbook (Beta v3) [Internet]. Singapore Government Developer Portal. 2023 [cited 24 Sept 2025].

<https://www.developer.tech.gov.sg/products/collections/data-science-and-artificial-intelligence/playbooks/prompt-engineering-playbook-beta-v3.pdf>.
