## Appendix D for "Evaluating Eight Retrieval-Augmented Generation (RAG) Large Language Models’ Responses to Clinical Questions: A Comparative Study"

### Appendix D. Critical key unique medical concepts per question

Treatment questions:

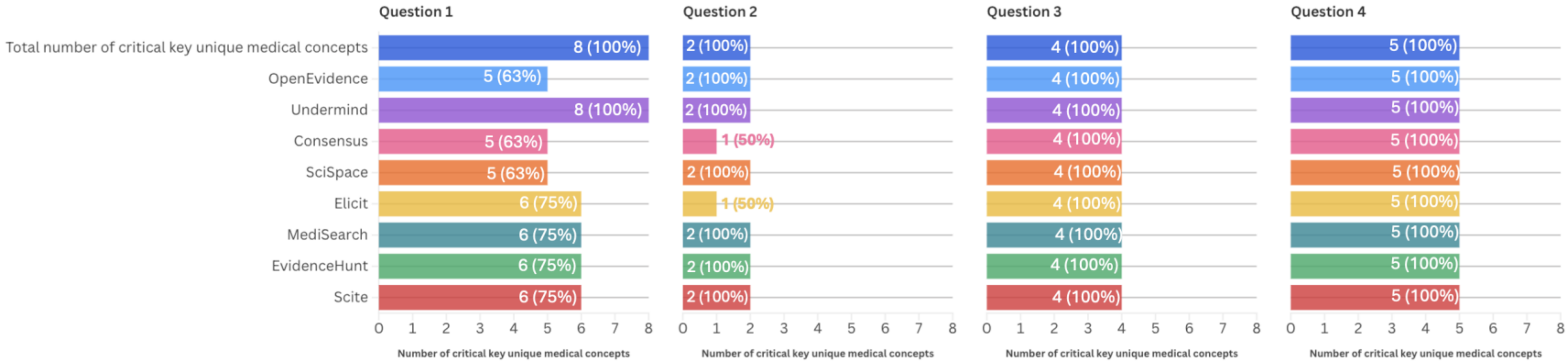


Etiology questions:

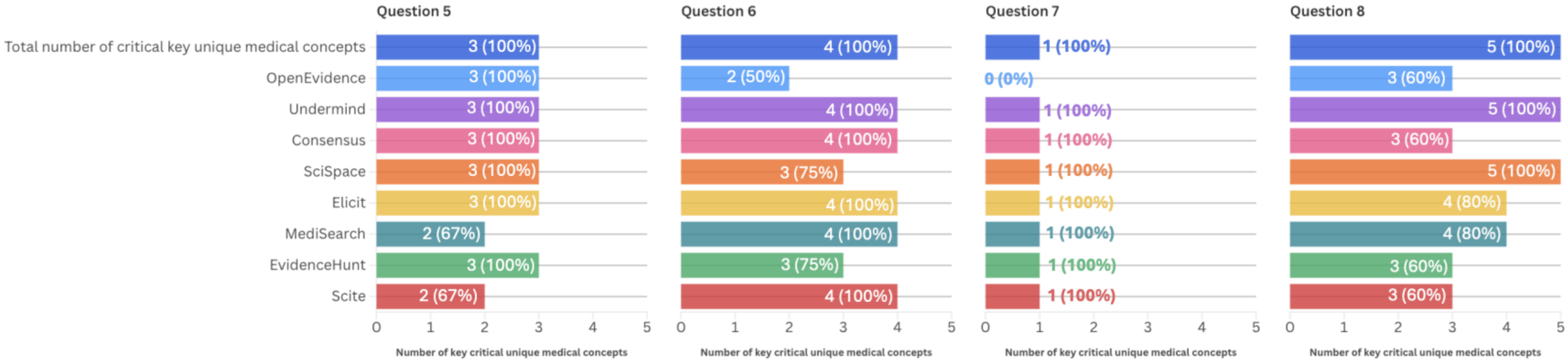

Prognosis questions:

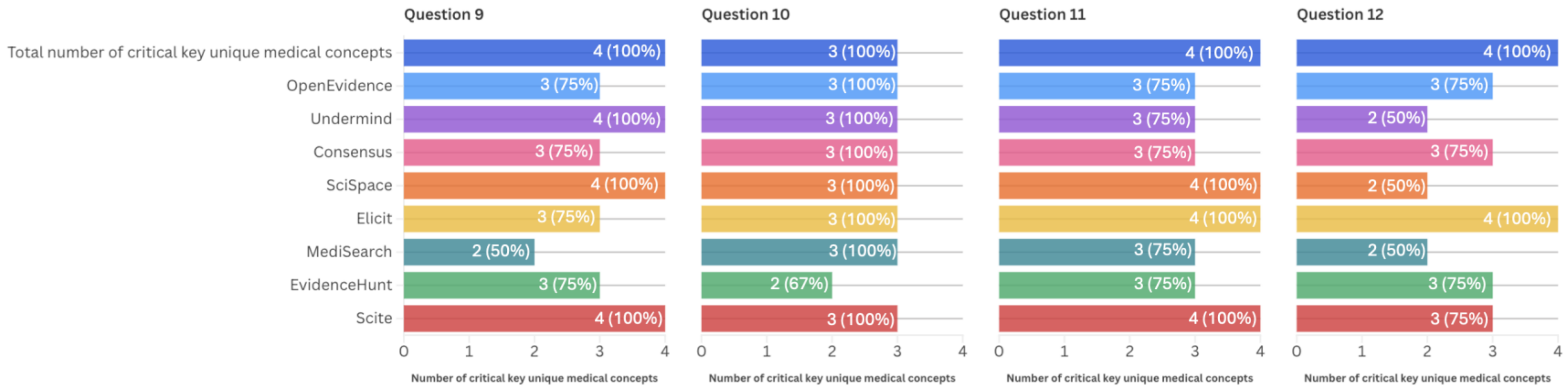


List of questions:

**Question 1:** In adults hospitalized with community-acquired pneumonia, does adjunctive corticosteroid therapy improve clinical recovery compared to standard antibiotic treatment alone?
